# The evidence on transmission dynamics of COVID-19 from pre- and asymptomatic cases: protocol for a systematic review (Version 2)

**DOI:** 10.1101/2021.05.06.21256615

**Authors:** T Jefferson, A Plüddemann, EA Spencer, J Brassey, EC Rosca, I Onakpoya, DH Evans, JM Conly, C Heneghan

**Affiliations:** The University of Oxford; Trip Database Ltd. https://www.tripdatabase.com; Victor Babes University of Medicine and Pharmacy of Timisoara; Li Ka Shing Institute of Virology and Dept. of Medical Microbiology & Immunology, University of Alberta; University of Calgary and Alberta Health Services, Calgary, Canada

**Keywords:** COVID-19, SARS-CoV-2, Transmission, Levels of evidence

## Abstract

**Background:** The role of forward transmission of infection from cases of SARS-CoV-2 who remain without symptoms and signs throughout the active phase of the disease (asymptomatic) and those who have not developed symptoms or signs when surveyed (presymptomatic) is at present unclear, despite the important role that they may play in transmission dynamics.

**Methods:** We will search LitCovid, medRxiv, Google Scholar and the WHO Covid-19 database using Covid-19, SARS-CoV-2, transmission, and appropriate synonyms as search terms. We will also search the reference lists of included studies for additional relevant studies. We will include studies of people exposed to SARS-CoV-2 within 2-14 days of close contact or suspected community or institutional exposure to index asymptomatic or pre-symptomatic infected individuals, as defined in each study along with secondary case(s) infected. We will include only studies that provide proof of transmission outcome using culturable virus and /or genetic sequencing. The inclusion of this higher-quality evidence aims to overcome the methodological shortcomings of lower quality studies. We will assess the microbiologic or genetic sequencing evidence in an effort to inform the quality of the chain of transmission evidence and adequacy of follow up of sign and symptom monitoring.

**Expected results:** We intend to present the evidence in three distinct packages: study description, methodological quality assessment and data extracted. We intend on summarising the evidence and drawing conclusions as to the quality of the evidence.

## Background

The overarching aim of the WHO’s <u>Global Strategic Preparedness and Response Plan for COVID-19</u> is to prevent transmission of SARS-CoV-2 and prevent associated illness and death. However, the transmission of the SARS-CoV-2 virus and the disease it causes is not completely understood, and public health and social measures (PHSMs) for restricting transmission are based on limited data with relatively few high-quality systematic reviews on the transmission of the SARS-CoV-2 virus available. To date, systematic reviews have revealed considerable methodological shortcomings in the included studies that hinder the development of firm conclusions over the transmission dynamics.^1-3^

Several reviews have addressed the magnitude of asymptomatic COVID-19 cases but the design and reporting of the included studies has revealed several deficits and biases, which have impacted the asymptomatic estimates and highlighted the need for more robust evidence^4^. Limitations identified have included the reliance on binary PCR testing alone to estimate asymptomatic fractions and onward transmission^5, 6^. Previous estimates of the asymptomatic influenza fraction are similarly affected by low quality study designs and methods. The role of cases which remain without symptoms or signs throughout the active phase (asymptomatic) of illness and those who have not developed symptoms or signs yet when surveyed (pre-symptomatic) is at present unclear, partly because of shortcomings in the methodologies employed in the studies^4, 7^.

A lack of standardised methods requires the integration of clinical, epidemiologic, molecular and laboratory evidence into a framework that identifies higher-quality evidence that reduces the uncertainty over the transmission dynamics of acute respiratory pathogens. The framework requires studies that use comprehensive and serial screening for symptoms^4^ and the use of high level confirmatory evidence of infection including viral culture and/or whole-genome sequencing to indicate the presence of replicable, infectious virus or with confirmation of identical sequences^8^.

This framework is a work in progress as scientific understanding in this area evolves.

### Objectives

To provide a rapid summary and evaluation of relevant data on the transmission of SARS-CoV-2 from pre and asymptomatic individuals, report important policy implications, and highlight research gaps of the highest priority.

## Methods

This review is part of a series of living reviews ^1-3, 9^ updated as new and important research is published. We set out to address the following questions:

1. Are asymptomatic or presymptomatic PCR positive individuals infectious;
2. What is the relationship between infectiousness and PCR cycle threshold;
3. If asymptomatic PCR positive individuals are infectious, what proportion are infectious and what is the duration of infectiousness;
4. Is there evidence of a chain of transmission that establishes asymptomatic and/or presymptomatic transmission of SARs-CoV-2?

### Search Strategy

The following electronic databases will be searched: LitCovid, medRxiv, Google Scholar and the WHO Covid-19 database. Search terms are COVID-19, SARS-CoV-2, transmission, and appropriate synonyms. The reference lists of included studies will be searched for additional relevant studies.

### WHO Covid-19 Database (https://search.bvsalud.org/global-literature-on-novel-coronavirus-2019-ncov/)

The global literature cited in the WHO COVID-19 database is updated daily (Monday through Friday) from searches of bibliographic databases, hand searching, and the addition of other expert-referred scientific articles.

### LitCovid (https://www.ncbi.nlm.nih.gov/research/coronavirus/)

A curated literature hub for tracking up-to-date scientific information about the 2019 novel Coronavirus. It is a comprehensive resource on the subject, providing central access to relevant articles in PubMed.

### medRxiv (https://www.medrxiv.org/)

A free online archive and distribution server for complete but unpublished manuscripts (preprints) in the medical, clinical, and related health sciences.

### Google Scholar (https://scholar.google.com/)

Provides a broad search for scholarly literature across many disciplines and sources: articles, theses, books, abstracts, from academic publishers, professional societies, online repositories, universities and other websites.

We will also search the bibliographies of retrieved systematic reviews^7^.

### Inclusion criteria

We will include studies if they reported the following information:

#### Population

people exposed to SARS CoV-2 within 2-14 days (incubation time) of close contact or suspected community or institutional exposure to index asymptomatic (at the time of observation) infected individuals, as defined in the study.

#### Reference

secondary case infected based on fulfilling a confirmed or probable case definition Target: level 3 / level 4 evidence with confirmed transmission outcome^8^.

#### Design

prospective or retrospective observational studies, including case series and ecological designs, or interventional studies including randomised trials and clinical reports, outbreak reports, case-control studies and experimental studies. Studies incorporating models to describe observed data will be included, however studies reporting solely predictive modelling will be excluded. Single case reports will be excluded, as no case report would have information on secondary cases.

Studies not reporting data by symptom status will be excluded as are those with a single observation point. A single observation or inadequate follow up cannot distinguish between presymptomatic, symptomatic and asymptomatic cases.

We modified our original protocol to define two <u>steps</u> of contribution to our evidence base from included studies. In the first <u>step,</u> we included all studies satisfying our overall inclusion criteria. In the second <u>step,</u> for assessing the chain of transmission (question 4), we will include those studies which identified index cases and reported sufficient data to document secondary transmission and thus allow analysis.

Sufficient data was defined as follows: i) documentation of transmission; ii) presence of replicating virus and/or documentation of phylodynamics (i.e. genetic sequence lineage); and iii) adequate follow-up and reporting of symptoms and signs. [See box 1]. For relevant studies, where necessary, one review author will write to the study corresponding author (with one reminder) to request further details.

**Box 1**

**i. Was transmission documented?**

**ia**. Was the chain of transmission adequately described and reported

Demonstrable and replicable chain of transmission (Gwaltney’s postulates^10^ and study replicability).

- Viral growth at the proposed anatomic site of origin;
- Culturable virus present in secretions or tissues shed from the site of origin;
- Infectious virus contamination and survival in or on environmental substrate or object;
- Viral contaminant reaches portal of entry of new host;
- The results of the study are replicated independently on the basis of the methods detailed in the first study.

The last item should be considered an ideal aim, as many transmission studies are one-off and observational. However, the body of high-quality evidence should be compatible with the conclusions on the mode of transmission. Outlying studies and those that reach different conclusions, should be assessed to ascertain reasons for diversity.

**ib**. Are the circumstances of transmission adequately assessed and reported?

- Context (exposure takes place).
- Environment (temperature, relative humidity, air exchanges, UV light etc).
- Route if known - report if multiple possible routes are entertained or cannot be ruled out.
- Circumstances of exposure, sample collection and signs and symptoms onset are recorded and reported.

**ii. Were viable replicating viruses and/or phylodynamics documented?**

**iia**. Presence of a viable replicating virus with phylodynamics compatible with hypothesised source ascertained

- Cq, Ct, Log concentration or number of copies are assessed and reported.
- Observed structural changes in host cells caused by viral invasion that leads to visible cell lysis and/or other cytopathic phenomena or equivalent in culture.
- Evidence of virus replication consistent with expected growth kinetics in appropriate cell lines.
- Testing for evidence of contamination by other infectious agents.

**iib** Serial Culture adequately described and reported

- Techniques measuring viral infectivity using appropriate cell lines (e.g. viral plaque assay, TCID50 and immunofluorescence).
- For guidance see section 11-b of *Use of cell culture in virology for developing countries in the South-East Asia Region. New Delhi: World Health Organization, Regional Office for South-East Asia; 2. Licence: CC BY-NC-SA 3*.*0 IGO*.

**iic** Genome sequencing adequately described and reported [based on <u>WHO Genomic</u> sequencing of SARS-CoV-2^11^ : items]

- Genome sampling strategies and study design are considered and reported, including the risk of cross-contamination [item 6.1].
- Appropriate metadata was collected and reported [item 6.2].
- The location of sequencing was appropriate [item 6.3.1].

iii. Was there a precise definition of symptoms and signs used and was follow up adequate?

Could the patient flow or data collection methods have introduced bias [based on QUADAS -2] and were measures to mitigate the bias introduced? ^12^

- A follow-up period is required to assess the presence or absence of symptoms and signs.
- Inadequate follow-up may misclassify pre-symptomatic individuals.
- An assessment of other underlying reasons for the presence of symptoms and signs should be applied in all cases

A reassessment of symptoms and signs should be recorded by another interviewer in a proportion of the cases as a data quality check.

### Quality Assessment

There are no formal quality assessment and reporting criteria for transmission studies, a situation reminiscent of the early days of Evidence-Based Medicine. Some authors have adapted observational checklists to assess quality^7^. However pre-existing tools and adaptations do not adequately account for the biases that might influence the understanding of the chain of transmission and the need to obtain microbiological as well as clinical confirmation of transmission. This is even more important in the case of asymptomatic transmission. This is why we have created the list in Box 1 and further broke down the contribution of each study into two steps and included only level 3 / level 4 evidence^8^.

Two reviewers will each extract data from the included primary studies and independently verify the data extraction of the other studies. Disagreements will be resolved by consensus or discussion with a third reviewer. One reviewer will assess the risk of bias from the primary studies and these judgments will be independently verified by a second reviewer.

### Data extraction

Search yields will be screened in duplicate and included study data will be extracted into templates that include study characteristics and methodological quality of studies and a summary of the main findings. References will be included in alphabetical order as a web appendix that facilitates updating. We will follow PRISMA reporting guidelines as indicated for systematic or scoping reviews where applicable (<u>PRISMA checklist</u>)^11^. Data extraction will be performed by one author and independently checked by a second author. Where there is disagreement, a third author will arbitrate.

### Data synthesis and reporting

Outcomes of interest are listed in the inclusion criteria. We will summarise data narratively and report the outcomes as stated in the paper, including quantitative estimates where feasible and relevant. We will also report subgroups of results by age categories where appropriate (e.g., school children, older adults). Where possible, compatible datasets of cycle threshold values and their relationship to viable replicating viruses will be analysed using appropriate statistical packages. Where necessary we will write to study authors for clarification of data. Where appropriate we will also report research and policy implications.

### Continual data release

As important new data accumulates, we will produce a report as an individual rapid review and aim to make all our work available by depositing the review findings in an open access repository (e.g. the <u>Oxford Research Archive</u>).

## Data Availability

All data included in the review will be provided in the tables and text

## Funding

This work is at present unfunded.

## Authors’ contributions

All authors contributed in equal part to the conceptualisation and development of the content. TJ and CH wrote the first draft and edited this version. All authors contributed to the subsequent drafts and approved the final version.

## Conflict of interest statements

TJ was in receipt of a Cochrane Methods Innovations Fund grant to develop guidance on the use of regulatory data in Cochrane reviews (2015 to 2018). In 2014 to 2016, he was a member of three advisory boards for Boehringer Ingelheim. TJ was a member of an independent data monitoring committee for a Sanofi Pasteur clinical trial on an influenza vaccine. TJ is occasionally interviewed by market research companies about phase I or II pharmaceutical products for which he receives fees (current). TJ was a member of three advisory boards for Boehringer Ingelheim (2014 to 16). TJ was a member of an independent data monitoring committee for a Sanofi Pasteur clinical trial on an influenza vaccine (2015 to 2017). TJ is a relator in a False Claims Act lawsuit on behalf of the United States that involves sales of Tamiflu for pandemic stockpiling. If resolved in the United States favour, he would be entitled to a percentage of the recovery. TJ is coholder of a Laura and John Arnold Foundation grant for the development of a RIAT support centre (2017 to 2020) and Jean Monnet Network Grant, 2017 to 2020 for The Jean Monnet Health Law and Policy Network. TJ is an unpaid collaborator to the project Beyond Transparency in Pharmaceutical Research and Regulation led by Dalhousie University and funded by the Canadian Institutes of Health Research (2018 to 2022). TJ consulted for Illumina LLC on next-generation gene sequencing (2019 to 2020). TJ was the consultant scientific coordinator for the HTA Medical Technology programme of the Agenzia per i Servizi Sanitari Nazionali (AGENAS) of the Italian MoH (2007 to 2019). TJ is Director Medical Affairs for BC Solutions, a market access company for medical devices in Europe. TJ was funded by NIHR UK and the World Health Organization (WHO) to update Cochrane review A122, Physical Interventions to interrupt the spread of respiratory viruses. TJ is funded by Oxford University to carry out a living review on the transmission epidemiology of COVID 19. Since 2020, TJ receives fees for articles published by The Spectator and other media outlets. TJ is part of a review group carrying out a Living rapid literature review on the modes of transmission of SARS CoV 2 (WHO Registration 2020/1077093 0). He is a member of the WHO COVID 19 Infection Prevention and Control Research Working Group for which he receives no funds. TJ is funded to co-author rapid reviews on the impact of Covid restrictions by the Collateral Global Organisation.

CJH holds grant funding from the NIHR, the NIHR School of Primary Care Research, the NIHR BRC Oxford and the World Health Organization for a series of Living rapid reviews on the modes of transmission of SARs CoV 2 reference WHO registration No2020/1077093. He has received financial remuneration from an asbestos case and given legal advice on mesh and hormone pregnancy tests cases. He has received expenses and fees for his media work including occasional payments from BBC Radio 4 Inside Health and The Spectator. He receives expenses for teaching EBM and is also paid for his GP work in NHS out of hours (contract Oxford Health NHS Foundation Trust). He has also received income from the publication of a series of toolkit books and for appraising treatment recommendations in non-NHS settings. He is the Director of CEBM, an NIHR Senior Investigator and an advisor to Collateral Global.

DE holds grant funding from the Canadian Institutes for Health Research and Li Ka Shing Institute of Virology relating to the development of Covid 19 vaccines as well as the Canadian Natural Science and Engineering Research Council concerning Covid 19 aerosol transmission. He is a recipient of World Health Organization and Province of Alberta funding which supports the provision of BSL3 based SARS CoV 2 culture services to regional investigators. He also holds public and private sector contract funding relating to the development of poxvirus based Covid 19 vaccines, SARS CoV 2 inactivation technologies, and serum neutralization testing.

JMC holds grants from the Canadian Institutes for Health Research on acute and primary care preparedness for COVID 19 in Alberta, Canada and was the primary local Investigator for a Staphylococcus aureus vaccine study funded by Pfizer for which all funding was provided only to the University of Calgary. He is a co-investigator on a WHO funded study using integrated human factors and ethnography approaches to identify and scale innovative IPC guidance implementation supports in primary care with a focus on low resource settings and using drone aerial systems to deliver medical supplies and PPE to remote First Nations communities during the COVID 19 pandemic. He also received support from the Centers for Disease Control and Prevention (CDC) to attend an Infection Control Think Tank Meeting. He is a member of the WHO Infection Prevention and Control Research and Development Expert Group for COVID 19 and the WHO Health Emergencies Programme (WHE) Ad hoc COVID 19 IPC Guidance Development Group, both of which provide multidisciplinary advice to the WHO, for which no funding is received and from which no funding recommendations are made for any WHO contracts or grants. He is also a member of the Cochrane Acute Respiratory Infections Group.

JB is a major shareholder in the Trip Database search engine (www.tripdatabase.com) as well as being an employee. In relation to this work, Trip has worked with a large number of organisations over the years, none have any links with this work. The main current projects are with AXA and Collateral Global.

ECR was a member of the European Federation of Neurological Societies(EFNS) / European Academy of Neurology (EAN) Scientist Panel, Subcommittee of Infectious Diseases (2013 to 2017). Since 2021, she is a member of the International Parkinson and Movement Disorder Society (MDS) Multiple System Atrophy Study Group, the Mild Cognitive Impairment in Parkinson Disease Study Group, and the Infection Related Movement Disorders Study Group. She was an External Expert and sometimes Rapporteur for COST proposals (2013, 2016, 2017, 2018, 2019) for Neurology projects.

IJO, EAS, and AP have no interests to disclose.

## Ethics committee approval

No approval was necessary

## Data Availability

All data included in the review will be provided in the tables and text.

## Notes

### Summary of Updates

Quality assessment and evidence synthesis sections have been updated

